# The First Large-Scale Wastewater Surveillance of AMR to inform Hyperlocal Antibiotic prescribing: A Study from Bengaluru, India

**DOI:** 10.1101/2024.08.15.24311496

**Authors:** Imon Roy, N Gowthami, Shoa Shamsi, Nikhilesh Unnikrishnan, Prerak Shah, Ramakrishna Prasad, Maneesh Paul Satyaseela, Varsha Shridhar

## Abstract

Approximately 1.2 million people died in 2019 globally due to antimicrobial resistant infections making it one of the major public health issues of the 21st century. Community level antibiotic resistance data is needed for guiding antibiotic prescriptions at the local level, which national databases may be unable to provide. We hypothesized that wastewater could have potential to detect local AMR patterns cost-effectively on a real time and longitudinal basis.

Our objectives were to explore the potential of wastewater to detect local AMR patterns of specific Gram Negative bacteria against 4 key antimicrobial classes across Bengaluru city. Wastewater from 66 open storm drain sites within all eight administrative zones of Bengaluru city, in southern India, between March and April 2022 were plated on McConkey and Shigella-Salmonella agar plates. 2 colonies were randomly picked from each plate, identified using standard biochemical tests and then subcultured for antibiotic sensitivity profiles against 5 common antibiotics: ciprofloxacin (Fluoroquinolone), cefixime (3rd generation cephalosporin), piperacillin/tazobactam (TZP, beta lactam/beta lactamase inhibitor), ertapenem and meropenem (carbapenems) on Mueller-Hinton plates

Of the 160 isolates selected for deeper scrutiny, 82% (132) belonged to the Enterobacteriaceae family. Of these, 95% (126) were known to cause human infections. About 28% of the clinically relevant Enterobacteriaceae isolates were susceptible to all five antibiotics tested while 12% were resistant to all five antibiotics, and the remainder had a mixed resistance profile. Our results indicate: a) drug resistant bacteria are found in significant numbers in open drains b) there is geographical variation within a city in the patterns and burden of AMR c) Wastewater could be both a method for estimating community burden of AMR. This is the first city-wide wastewater based AMR profiling in India and indicates the potential of such surveillance to provide hyperlocal community level data to influence local antibiotic prescription practices by primary care physicians and on antibiotic purchase and allocation decisions by government drug purchase committees.

## Introduction

Approximately 1.2 million people died in 2019 globally due to antimicrobial resistant infections making it one of the major public health issues of the 21st century (1). Yet, antimicrobial resistance, contrary to popular belief, is not just a problem created in the white rooms of doctors’ clinics and under the harsh lights of the ICUs. It is a “wicked” problem spanning human, animal and environmental realms (2,3), which epitomizes volatility, uncertainty, complexity and ambiguity and the need for nuanced understanding and interpretation-all the characteristics that make it incredibly challenging to solve through broad public health directives or vertical programs.

### The Need for Community-Level Antimicrobial Resistance and the Potential of Wastewater Testing

Antimicrobial resistance both contributes to and is heavily influenced by local factors at the community and the individual levels (4). Antimicrobial stewardship, the process of fighting antibiotic resistance and preserving antibiotics, is, at its foundation, a behavioral change at the level of individuals and local organizations. Behavioral changes need the problem to be personalized, for individuals to believe in a sense of personal urgency without which it is impossible to keep efforts sustained for altering lifelong habits.

However, currently, there is a perceptible translation gap between the data and directives of antimicrobial resistance and the day to day practice of antibiotic prescribing. Without a sense of personal agency or responsibility, antimicrobial stewardship on the ground remains theoretical. This could be due to the fact that much of the data on AMR is derived from clinical specimens received at large, tertiary care hospitals attached to clinical academic research centers, thus increasing the chances that it is seen as irrelevant at the ground level. Thus, while national databases and trend reports for antibiotic resistance have their place and uses, there are dangers in using them as guidelines on local prescriptions.

### Wastewater as a Surrogate for Measuring Community Burden of AMR

Community level antibiotic resistance data is notoriously difficult to obtain. Testing individual clinical samples from communities for the purpose of surveillance is labour-intensive and expensive, not to mention mired in ethical dilemmas about privacy and availability/ accessibility of subsequent treatment. While each hospital is expected to generate its own antibiograms (which track antibiotic usage and antimicrobial resistance against the antibiotics), which may or may not be regularly updated (5), this data is usually not shared publicly or entered into a central database for policy analysis and surveillance. How then to generate local community-relevant, yet anonymized, data that could enable bottom-up, local-context based initiatives which are driven by motivations that arise not just from the cognitive domain but also in the affective domain?

We believe that wastewater could be such a medium. Wastewater surveillance had been used as far back as 1900 to track *Mycobacterium tuberculosis* from sanatoriums (6), in the 1930s to detect typhoid outbreaks in villages (7), is now most well-known for usage as a monitoring method for polio virus (8) and in the recent past has seen a revival as an early warning system for Covid-19 (9,10).

Wastewater is a significant environmental reservoir of AMR. It could also contain high levels of antibiotics from various sources (Antibiotic manufacturers, human, veterinary, surface runoff from animal husbandry industries etc) and thus represents an ideal environment for various populations of susceptible and resistant bacteria (ARB) to co-exist, exchange antimicrobial resistant genes (ARGs) and amplify and spread AMR (11). Many studies in the past decade have shown the presence of ARGs against beta lactams (12) fluoroquinolones (12), cephalosporins, colistin, carbapenems (13) and more, using methods ranging from Kirby Bauer method, PCR and metagenomic approaches (11). AMR reservoirs in the environment could be a driver of persistence of AMR, even in countries with strong antimicrobial stewardship. Microbes can acquire antibiotic resistant genes from each other between human, animal and environmental domains.

In this paper, our objectives were to explore the potential of wastewater to detect local AMR patterns by characterizing the phenotypic patterns of specific Gram Negative Bacteria (GNB) against 4 key antimicrobial classes and 5 commonly used exemplar antibiotics across Bengaluru city. We created a hyperlocal AMR map of Bengaluru city showing the patterns and burden of resistance and we highlight the potential of using such maps, when regularly updated and communicated to primary care physicians of the same localities, to inform about local AMR patterns and influence antibiotic prescription.

## Methods

### Sampling sites

Bengaluru city is India’s third most populous city, with a total population of about 13.1 million individuals, spread over 2000 square kilometers (14). About 60-70% of its wastewater is underground and part of a formal sewage network, while 30% is disposed through open storm water drains. 66 open drain sites belonging to 8 municipal administrative zones, as designated by the Municipal Corporation of Bengaluru city (Figure 1), were sampled over a period of 2 months from March to April 2022. The average temperature in Bengaluru during this time of the year is between 23-26^°^C. Of the 66 sites sampled, 14 were sampled in Mar 22 and the remaining in April 22.

**Figure 1.**
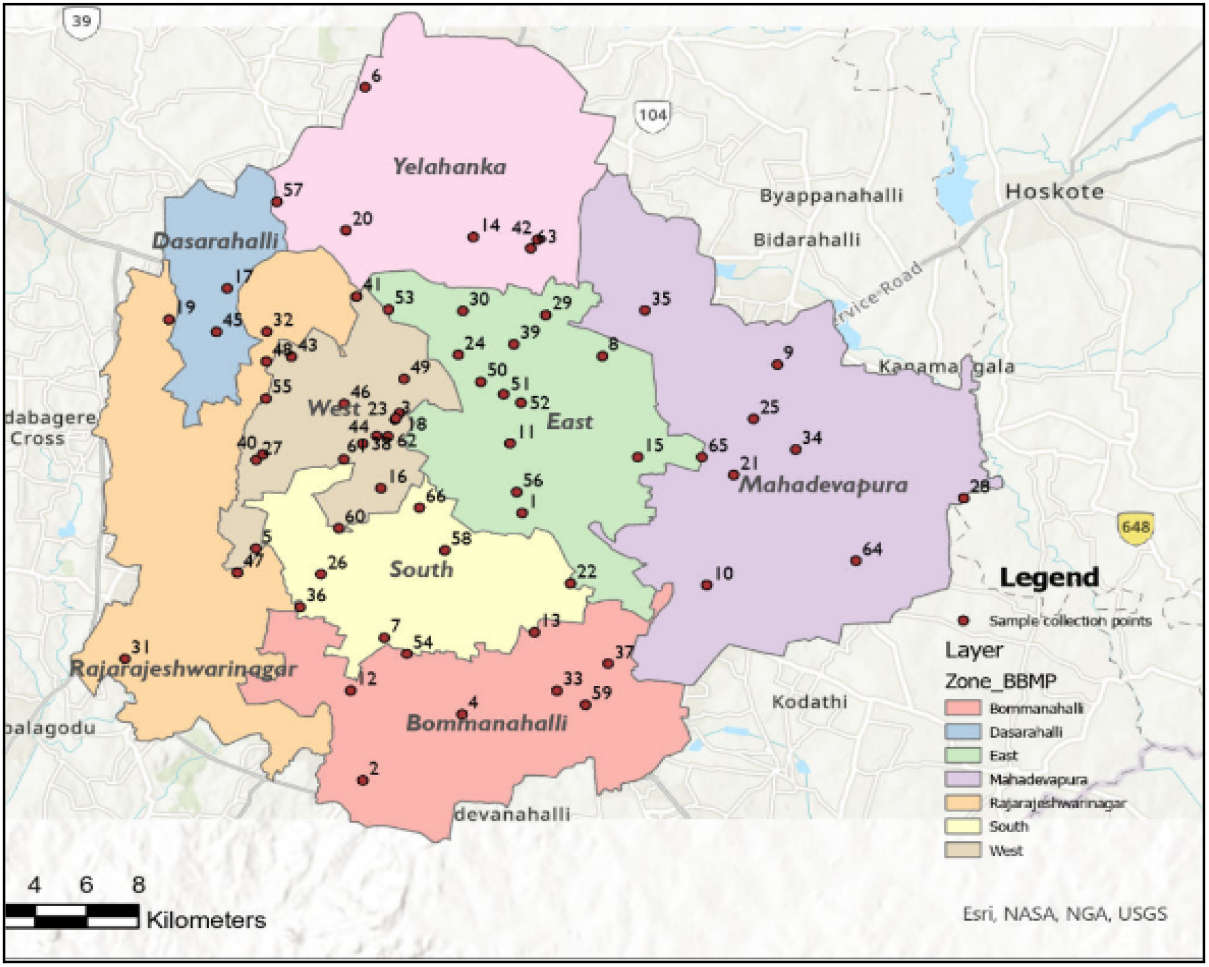
Map of Bangalore, showing the administrative zones and the collection sites

### Sample collection

About 500mL of wastewater samples from open drains were collected in clean low density polyethylene bottles using the grab and collect method. pH was measured at the time of collection and, if found to be between 7 and 7.2, the samples were transported to the lab on ice packs, within one to two hours of collection. Samples were plated as described in the section below.

### Microbiological culture

Two different selective media plates were used for this pilot - MacConkey agar and Salmonella-Shigella (SS) agar. Our target was to isolate Gram negative bacteria from MacConkey &Salmonella-Shigella agar plates. Fresh wastewater sample was serially diluted to 1:1000 using 0.9% saline. From the diluted sample, 50µl of sample was plated onto the Mac Conkey plate and SS agar plate separately. Samples were spread evenly using sterile L shaped rod and incubated overnight. Colonies were counted and single colonies were randomly picked for further analysis from the plates, subcultured and identified using Himedia Biochemical strip test kits [HiAssorted™ Biochemical Test Kit and HiIMViC™ Biochemical Test Kit]. These colonies were grouped in two different ways: A) Clinically Relevant or Environmental (based on published literature on where it is commonly isolated from) and b) Enterobacteriaceae or Non - Enterobacteriaceae.

### Sensitivity assays

For antimicrobial resistance, Kirby Bauer disc diffusion method was followed. 100µl of the subculture was added onto Mueller Hinton Agar (MHA) plates and evenly spread on the media plate and dried. Five antibiotics discs at standard concentrations-Meropenem 10mcg (SD727), Ertapenem 10mcg (SD280), Ciprofloxacin 5mcg (SD060), Cefixime 5mcg (SD266) and Piperacillin/ Tazobactam 100/10mcg (SD210) (Himedia Labs) were placed on the MHA plates equally spaced and the plates were incubated overnight. The zone of inhibition was calculated by measuring the distance from the center of the disk to the farther end of the inhibition zone. These measurements were taken by two individuals. The inhibition zones were measured to determine resistance or susceptibility of each isolate to the antibiotics according to the Clinical and Laboratory Standards Institute (CLSI) standards (Supplementary table 3).

The isolates were mapped into their respective municipal zones. Clinically relevant Enterobacteriaceae family isolates were summarized into those that are resistant against only ciprofloxacin, cefixime, TZP or both carbapenems, then a combination of antibiotic pairs, and antibiotic triads (Table 1). For the purposes of this summary, intermediate resistance was considered to be susceptible as the antibiotic in question still acts against the pathogen, only at higher concentrations.

**Table 1:** Zone wise resistance profiles of all clinically relevant Enterobacteriaceae isolates Legends – MER-Meropenem; ERT – Ertapenem; CIP – Ciprofloxacin; CF – Cefixime; TZP – Piperacillin/Tazobactam.

| Zone | Number of isolates | Susceptible % (n) | Resistant only to CIP | Resistant only to CF | Resistant only to TZP | Resistant only to both ERT and MER | Resistant to ERT or MER | Resistant only to CF and CIP | Resistant to CF, CIP and TZP | Resistant to TZP and ERT & MER only | Resistance | Other combinations |
| --- | --- | --- | --- | --- | --- | --- | --- | --- | --- | --- | --- | --- |
| West | 30 | 38% (11) | 3.4% (1) | 6.8% (2) | 3.4% (1) | 0 | 20.4% (6) | 6.8% (2) | 0 | 6.8% (2) | 6.8% (2) | 6.8% (2) |
| South | 19 | 21% (4) | 10.5% (2) | 0 | 0 | 0 | 10.5% (2) | 10.5% (2) | 0 | 15.8% (3) | 31.6% (6) | 0 |
| East | 27 | 40.7% (11) | 11.1 % (3) | 7.4% (2) | 0 | 3.7% (1) | 22.2% (6) | 0 | 3.7% (1) | 7.4% (2) | 0 | 3.7% (1) |
| Yelahanka | 8 | 12.5% (1) | 12.5% (1) | 0 | 0 | 12.5% (1) | 25% (2) | 0 | 0 | 12.5% (1) | 0 | 25% (2) |
| Mahadevpura | 18 | 27.8% (5) | 5.6% (1) | 11.1% (2) | 0 | 5.6% (1) | 11.1% (2) | 0 | 0 | 16.7% (3) | 16.7% (3) | 5.6% (1) |
| Rajarajeshwari Nagar | 10 | 10% (1) | 0 | 0 | 0 | 0 | 50% (5) | 0 | 0 | 0 | 10% (1) | 30% (3) |
| Bommanahalli | 11 | 18% (2) | 0 | 0 | 0 | 0 | 18% (2) | 9% (1) | 9% (1) | 0 | 27% (3) | 18% (2) |
| Dasarahalli | 6 | 16.7% (1) | 0 | 0 | 0 | 0 | 33.3% (2) | 0 | 0 | 0 | 16.7% (1) | 0 |
| Total | 128 | 28.1 % (36) | 6.3% (8) | 4.7% (6) | 0.8% (1) | 2.3% (3) | 21.1% (27) | 3.9% (5) | 1.6% (2) | 8.6% (11) | 12.5% (16) | 8.6% (11) |

### Data visualization

The data obtained in this study has been mapped and summarized here: https://storymaps.arcgis.com/stories/011a7411f6ed44c5840971cda1b6f0b9 for easy browsing.

## Results

### Data description

A total of 160 colonies were isolated during the study period, of which 28 were isolated in March 2022 and the remainder in April 2022. These 160 isolates fell into 29 species. 150 out of the 160 were considered clinically relevant, with 132 belonging to the Enterobacteriaceae family (Supplementary Tables 1 and 2). The most common species found were *Klebsiella pneumoniae* (18 *Klebsiella pneumoniae* subsp. *pneumoniae and 3 Klebsiella pneumoniae* subsp. *Ozaenae* isolates) and *Escherichia coli (*15 isolates).

### Significant presence of multi drug resistant bacteria in the wastewater system across all geographical zones in the city

We found that 28% of all clinical Enterobacteriaceae isolates were susceptible to all the tested antibiotics, whereas 12.5% were resistant to all the tested antibiotics. About 4% of all isolates were resistant only to Ciprofloxacin and cefixime, while 12% were resistant only to ciprofloxacin or cefixime or TZP; about 9% were resistant to the higher order antibiotics only (TZP and carbapenems). The remainder had a mixed combination of resistance profiles. Interestingly, about a fifth of isolates (21%) which were resistant to ertapenem were not found to be resistant to meropenem (Table 1).

We then classified all clinically relevant bacteria, Enterobacteriaceae and non-Enterobacteriaceae, by zone and number of isolates resistant to increasing numbers of antibiotics.

### Geographical variation within a city in the patterns and burden of AMR

Multidrug resistance, as defined as resistance against 3 or more classes of antibiotics (15), was found in all eight zones, at varying levels (Figure 2). The proportion of resistant organisms to each of the antibiotics tested varied between the 8 zones (Supplementary table 3). The number of organisms with resistances to 2 or more antibiotics also showed significant spatial variation (Fig 2).

**Figure 2.**
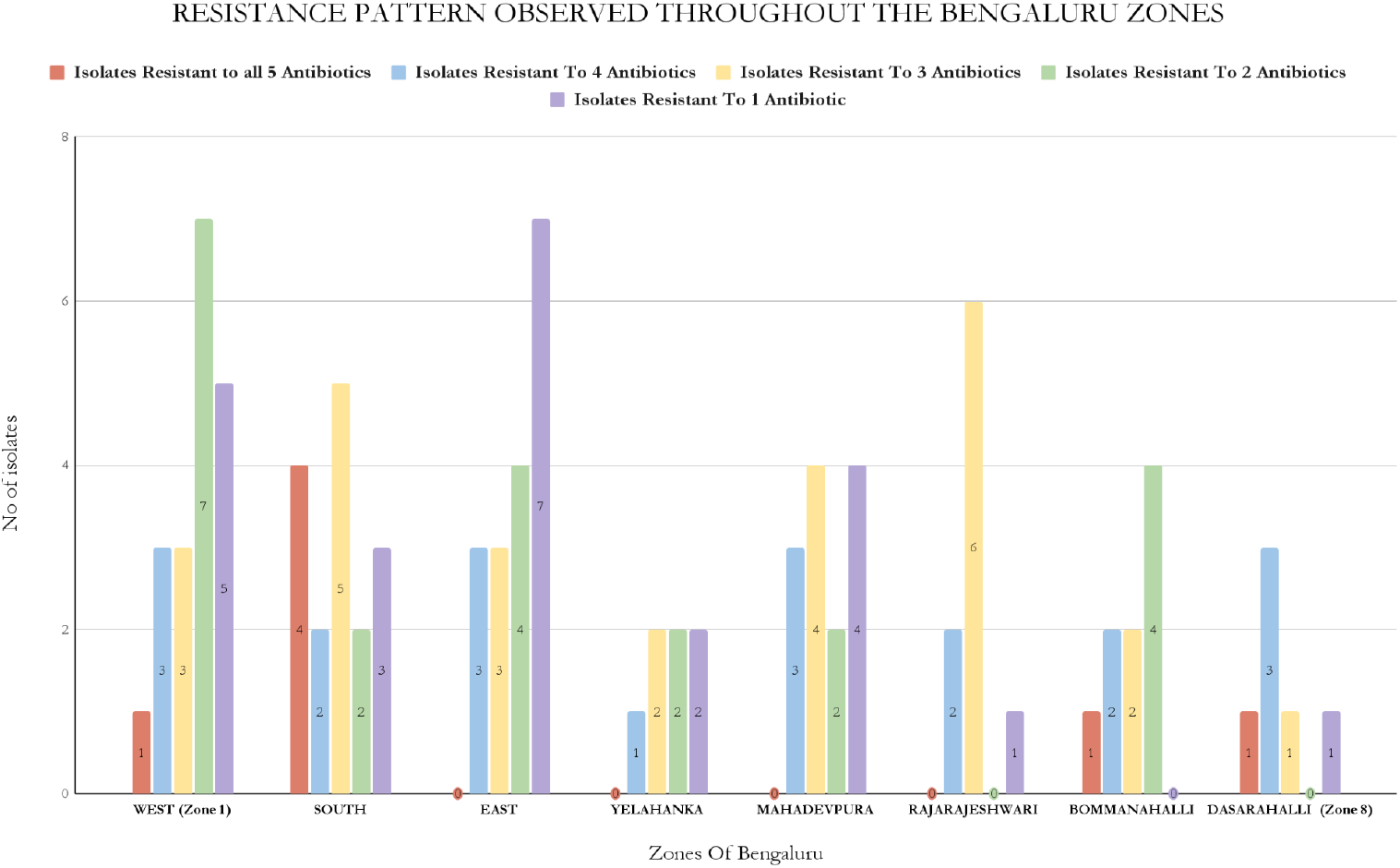
Resistance Pattern Observed Throughout The Bengaluru Zones

### Resistance patterns of *Klebsiella pneumoniae* and *Echerichia coli*

*Klebsiella pneumoniae* and *Echerichia coli* were the two most abundantly isolated species. They are also highly clinically relevant with significant resistance being reported in India and worldwide. Of the 21 isolates of *Klebsiella pneumoniae*, 18 were *Klebsiella pneumoniae* subspecies *pneumoniae* and the remaining were subspecies *ozaenae*. Of the 18, 7 (∼40%) showed complete (n=3) or intermediate (n=4) resistance to all antibiotics tested, and 2 were completely susceptible to all antibiotics and the remaining showed a combination of resistance profiles. Among the 15 isolates of E.coli, 3 (20%) showed complete resistance to all 5 antibiotics.

### AMR profiles of environmentally relevant samples

There was a single species of environmentally relevant organisms found: *Buttiauxella agrestis. Buttiauxella agrestis* was found to be susceptible to all antibiotics except cefixime, against which it had intermediate resistance (Supplementary Table 1).

## Discussion

To our knowledge, this is the first city-wide wastewater based AMR profiling in India and indicates the potential of such surveillance to provide hyperlocal community level data. This data can be used to influence local antibiotic prescription practices by primary care physicians, on the targeted sales of antibiotics by pharma companies and on antibiotic purchase and allocation decisions by drug purchase committees.

In this pilot study we investigated the pattern of AMR in the wastewater collected across the 8 administrative zones of Bengaluru city, Karnataka, India to five antibiotics belonging to four different classes - Meropenem and Ertapenem (Carbapenems), Piperacillin - Tazobactam (Penicillin + Beta lactamase inhibitor), Cefixime (Cephalosporin) and Ciprofloxacin (Fluoroquilones).

### Implications of finding drug resistant bacteria in significant numbers in open drains

Our study indicates a significant presence of multi drug resistant bacteria in the wastewater system. In Bengaluru, about 70% of the water in 2021 was estimated to flow in underground sewage pipes which are part of the formal sewerage network which undergoes treatment before being released into lakes and other reservoirs. Unlike other studies done on influent or effluent water from sewage treatment plants (STPs) (11,16), our study is unique in that the samples were collected from the residual 30% of wastewater flowing in open storm drains. This wastewater comes from slums and illegal pipelines from establishments that are not connected to the formal STP network. As such this water could provide a window into populations that may not access formal healthcare systems and are typically hidden from the mainstream. The presence of carbapenem resistance in this water could indicate the incorrect disposal of antibiotics, the non-linkage of hospitals to the formal sewerage network, or the inappropriate usage and disposal of higher level antibiotics in non-hospital settings.

Water from open storm drains usually empty into water bodies such as lakes without treatment. This water is then used for fishing, agricultural irrigation or pumped for use as drinking water within the city, or pumped to peri-urban and rural areas for the same purposes. Thus open drains represent a path by which AMR circulates and escalates within the city and from the city to its surroundings. They may indicate routes by which individuals who have not been exposed to high levels of antibiotics end up with resistant bacteria.

### Antibiotic Resistance profiles of *Klebsiella pneumoniae, Escherichia coli* isolates suggest multiple resistance mechanisms at play

*Klebsiella pneumoniae* and *Escherichia coli* are among the most common pathogens associated with severe multi-drug resistant nosocomial infections in India (17). They were also the most commonly isolated organisms in our study. Thus, they merited a closer examination of their resistance profiles.

Some unusual patterns noticed among the *Klebsiella pneumoniae* and *Escherichia coli* isolates and their possible reasons (barring occasional technical errors) are described below. Since resistance to ciprofloxacin is conferred by a separate set of genes encoding DNA gyrase, topoisomerase or the PMQR family (18), it is not discussed in further detail below.

Thus, our data indicates that resistance to one member of a class of antibiotics does not necessarily remove the entire class of antibiotics nor the classes below the antibiotic level from the armamentarium among isolates present in wastewater.

| Pattern | Possible reasons |
| --- | --- |
| Carbapenem Resistant.<br>TZP Susceptible<br>Cefixime Susceptible | Resistance mutations in efflux pumps or porins (for instance, mutations that change the charge of porins can affect piperacillin differently from meropenem or ertapenem (19) |
| Carbapenem Resistant.<br>TZP Resistant<br>Cefixime Susceptible | Presence of OXA-48 gene (20) preserves susceptibility to cephalosporins while compromising carbapenem and other beta lactam/lactamase activities |
| Carbapenem Resistant<br>TZP Susceptible<br>Cefixime Resistant | Mutations in porins could affect carbapenems without affecting TZP (19).<br>Presence of ESBLs affect 3rd generation cephalosporins like cefixime, but can be overcome by the presence of tazobactam in TZP which is a beta lactamase inhibitor |
| Carbapenem Sensitive<br>TZP Resistant<br>Cefixime Resistant | No presence of genes coding for carbapenemases. This phenotype could also be due to genes such as TEM 3-7, SHV or CTX-M family of genes which confer resistance to penicillins and cephalosporins (21). |
| Differential Resistance Profiles between Meropenem and Ertapenem | Nearly 20% of the isolates show differential resistance profiles between these two carbapenems. This discrepancy could be due to three reasons:<br>a) ertapenem is known to have a lower threshold for resistance than meropenem (22) |
|  | b) Ertapenem resistance is known to be enhanced by the presence of ESBL and point mutations in the porin genes to a much larger extent than meropenem (23)<br>c) The half life of ertapenem is 4 hours compared to meropenem's 1 hour (22), making it likely that the wastewater environment potentially contains more ertapenem than meropenem. |

### Limitations

This study did not delve into the antibiotic resistance genes causing these phenotypes which would be extremely important and essential in future iterations. We also were unable to test for the levels of active antibiotics in the wastewater samples, which would have given an indication of whether the AMR bacteria were being shed from certain super-generators (such as hospitals or poultry farms or animal husbandry units) or actively being selected for in an environment that had persistently high levels of antibiotics, as shown elsewhere (24). A major limitation is also that this study is cross-sectional. Overall resistances in a city can be obtained only when there are longitudinal trends and not from a single set of readings. The role or impact of bacteriophages was not determined in this study. Where bacteria thrive, so will bacteriophages, viruses that infect bacteria. Bacteriophages are considered to be agents of horizontal gene transfer through the process of transduction and thus, contributors to the spread of ARGs (25). They could also simultaneously be considered to play a beneficial role by controlling the population of AMR bacteria (26). Wastewater, fecal sludge-containing open drains etc are complex environments where bacteria and bacteriophages exist in the presence of antibiotics and other chemicals which can have numerous unknown and unpredictable effects on ARG transfer and the actual disease causation.

## Conclusions

This study investigates the antimicrobial resistance patterns of 4 groups of antibiotics throughout the different zones of Bengaluru city, Karnataka. Our results indicate: a) wastewater could be a novel and cost-effective method of determining hyperlocal AMR patterns and burden in communities b) drug resistant bacteria are found in significant numbers in open drains c) there is geographical variation within a city of the patterns and burden of AMR, opening up the possibility of community-based and community-relevant antimicrobial stewardship practices. Wastewater surveillance also allows for community-participation in surveillance and for grassroots or participatory initiatives towards all 6 pillars of the National Action Plan for Antimicrobial Resistance (27), contributing to increased awareness, strengthening knowledge/evidence base, promoting AMR control measures & optimized use of antimicrobials, facilitating novel avenues for investments in AMR research and innovations and finally, showcasing India’s commitment to and leadership in antimicrobial stewardship, in line with India’s position as President of the G20 with a mandate for the same.

## Data Availability

All data produced in the present study are available upon reasonable request to the authors.

https://storymaps.arcgis.com/stories/011a7411f6ed44c5840971cda1b6f0b9

