## Supplementary Table 1 for "The First Large-Scale Wastewater Surveillance of AMR to inform Hyperlocal Antibiotic prescribing: A Study from Bengaluru, India"

**Supplementary Table 1:** Isolates from all zones of Bengaluru City were tested for antibiotic susceptibility by Kirby Bauer method.

R -Resistance, IR - Intermediate resistance and S - Susceptible as per CLSI standards.

| **ZONE** |  | **BACTERIA IDENTIFIED** | **Zone of Inhibition** | | | | | | | | | |
| --- | --- | --- | --- | --- | --- | --- | --- | --- | --- | --- | --- | --- |
|  |  |  | **Meropenem** | | **Ertapenem** | | **Pippercillin/**  **Tazobactam** | | **Cefixime** | | **Ciprofloxacin** | |
|  |  |  | Zone inhibition calculated (mm) | Inference | Zone inhibition calculated (mm) | Inference | Zone inhibition calculated (mm) | Inference | Zone inhibition calculated (mm) | Inference | Zone inhibition calculated (mm) | Inference |
| **WEST** | **S. No** | **CLINICALLY RELEVANT** | | | | | | | | | | |
|  | 1 | *Escherichia coli* | 1 | R | 1 | R | 0 | R | 0 | R | 1 | R |
|  | 2 | *Kluyvera ascorbata* | 26 | S | 20 | IR | 26 | S | 10 | R | 30 | S |
|  | 3 | *Escherichia coli* | 28 | S | 19 | IR | 20 | IR | 6 | R | 1 | R |
|  | 4 | *Klebsiella pneumoniae subspecies pneumoniae* | 20 | IR | 18 | R | 16 | IR | 15 | R | 20 | R |
|  | 5 | *Kluyvera ascorbata* | 9 | R | 1 | R | 15 | R | 20 | IR | 19 | R |
|  | 6 | *Shigella species* | 50 | S | 50 | S | 30 | S | 35 | S | 26 | S |
|  | 7 | *Kluyvera ascorbata* | 21 | IR | 28 | S | 22 | S | 18 | IR | 32 | S |
|  | 8 | *Escherichia coli* | 26 | S | 22 | S | 20 | IR | 15 | R | 24 | S |
|  | 9 | *Salmonella choleraesuis subspecies choleraesuis* | 21 | S | 19 | IR | 16 | R | 20 | IR | 20 | R |
|  | 10 | *Escherichia hermanii* | 45 | S | 50 | S | 50 | S | 50 | S | 25 | S |
|  | 11 | *Kluyvera ascorbata* | 20 | IR | 13 | R | 30 | S | 32 | S | 24 | S |
|  | 12 | *Enterobacter amnigenus* | 39 | S | 38 | S | 40 | S | 37 | S | 30 | S |
|  | 13 | *Salmonella choleraesuis subspecies choleraesuis* | 19 | IR | 17 | R | 24 | S | 21 | S | 24 | S |
|  | 14 | *Klebsiella pneumoniae subspecies pneumoniae* | 23 | S | 19 | IR | 20 | IR | 19 | IR | 23 | S |
|  | 15 | *Serratia sp* | 15 | R | 8 | R | 17 | R | 26 | S | 35 | S |
|  | 16 | *Escherichia hermanii* | 29 | S | 10 | R | 1 | R | 1 | R | 20 | R |
|  | 17 | *Shigella flexneri* | 20 | IR | 21 | S | 21 | S | 8 | R | 20 | R |
|  | 18 | *Klebsiella pneumoniae subspecies pneumoniae* | 40 | S | 35 | S | 32 | S | 35 | S | 40 | S |
|  | 19 | *Proteus mirabilis* | 14 | R | 6 | R | 18 | IR | 9 | R | 23 | S |
|  | 20 | *Serratia sp* | 40 | S | 31 | S | 25 | S | 34 | S | 39 | S |
|  | 21 | *Citrobacter freundii* | 32 | S | 25 | S | 26 | S | 30 | S | 26 | S |
|  | 22 | *Klebsiella pneumoniae subspecies pneumoniae* | 20 | IR | 16 | R | 22 | S | 5 | R | 21 | R |
|  | 23 | *Serratia sp* | 25 | S | 18 | R | 15 | R | 1 | R | 8 | R |
|  | 24 | *Enterobacter aerogenes* | 46 | S | 39 | S | 40 | S | 45 | S | 30 | S |
|  | 25 | *Shigella flexneri* | 42 | S | 34 | S | 30 | S | 40 | S | 36 | S |
|  | 26 | *Enterobacter aerogenes* | 26 | S | 23 | S | 19 | IR | 23 | S | 13 | R |
|  | 27 | *Salmonella choleraesuis subspecies choleraesuis* | 26 | S | 16 | R | 25 | S | 15 | R | 25 | S |
|  | 28 | *Enterobacter amnigenus* | 40 | S | 41 | S | 43 | S | 35 | S | 36 | S |
|  | 29 | *Kluyvera ascorbata* | 20 | IR | 13 | R | 15 | R | 25 | S | 22 | S |
|  | 30 | *Serratia plymuthica* | 24 | S | 25 | S | 20 | IR | 15 | R | 20 | R |
| **SOUTH** | **S. No** | **CLINICALLY RELEVANT** | | | | | | | | | | |
|  | 1 | *Escherichia hermanii* | 20 | IR | 20 | IR | 0 | R | 15 | R | 1 | R |
|  | 2 | *Kluyvera ascorbata* | 19 | IR | 12 | R | 9 | R | 1 | R | 1 | R |
|  | 3 | *Escherichia coli* | 18 | R | 20 | IR | 15 | R | 14 | R | 18 | R |
|  | 4 | *Salmonella choleraesuis subspecies choleraesuis* | 23 | S | 20 | IR | 18 | IR | 15 | R | 20 | R |
|  | 5 | *Pantoea dispersa* | 39 | S | 35 | S | 39 | S | 40 | S | 50 | S |
|  | 6 | *Shigella species* | 20 | IR | 13 | R | 20 | IR | 24 | S | 35 | S |
|  | 7 | *Klebsiella pneumoniae subspecies pneumoniae* | 15 | R | 17 | R | 0 | R | 10 | R | 11 | R |
|  | 8 | *Pantoea dispersa* | 17 | R | 10 | R | 15 | R | 31 | S | 27 | S |
|  | 9 | *Serratia sp* | 20 | IR | 25 | S | 20 | IR | 21 | S | 20 | R |
|  | 10 | *Escherichia coli* | 18 | R | 6 | R | 14 | R | 6 | R | 19 | R |
|  | 11 | *Shigella flexneri* | 39 | S | 40 | S | 35 | S | 45 | S | 1 | R |
|  | 12 | *Escherichia coli* | 35 | S | 25 | S | 26 | S | 35 | S | 25 | S |
|  | 13 | *Serratia entomophila* | 22 | S | 16 | R | 26 | S | 26 | S | 16 | R |
|  | 14 | *Escherichia coli* | 9 | R | 1 | R | 12 | R | 28 | S | 26 | S |
|  | 15 | *Citrobacter freundii* | 26 | S | 26 | S | 26 | S | 30 | S | 27 | S |
|  | 16 | *Escherichia hermanii* | 1 | R | 1 | R | 1 | R | 1 | R | 1 | R |
|  | 17 | *Salmonella choleraesuis subspecies choleraesuis* | 35 | S | 26 | S | 34 | S | 26 | S | 28 | S |
|  | 18 | *Klebsiella pneumoniae subspecies pneumoniae* | 1 | R | 1 | R | 1 | R | 1 | R | 1 | R |
|  | 19 | *Shigella species* | 15 | R | 10 | R | 17 | R | 31 | S | 35 | S |
|  | 20 | *Serratia plymuthica* | 9 | R | 13 | R | 20 | IR | 30 | S | 17 | R |
| **EAST** | **S. No** | **CLINICALLY RELEVANT** | | | | | | | | | | |
|  | 1 | *Enterobacter aerogenes* | 24 | S | 15 | R | 24 | S | 13 | R | 25 | S |
|  | 2 | *Proteus mirabilis* | 35 | S | 29 | S | 31 | S | 31 | S | 26 | S |
|  | 3 | *Enterobacter amnigenus* | 16 | R | 1 | R | 13 | R | 25 | S | 20 | R |
|  | 4 | *Citrobacter freundii* | 25 | S | 14 | R | 15 | R | 25 | S | 13 | R |
|  | 5 | *Pantoea dispersa* | 35 | S | 30 | S | 30 | S | 27 | S | 28 | S |
|  | 6 | *Shigella species* | 50 | S | 50 | S | 50 | S | 50 | S | 50 | S |
|  | 7 | *Escherichia coli* | 50 | S | 50 | S | 50 | S | 50 | S | 50 | S |
|  | 8 | *Proteus vulgaris* | 23 | S | 22 | S | 24 | S | 19 | S | 20 | R |
|  | 9 | *Escherichia hermanii* | 40 | S | 29 | S | 24 | S | 35 | S | 27 | IR |
|  | 10 | *Shigella flexneri* | 16 | R | 0 | R | 19 | IR | 13 | R | 25 | IR |
|  | 11 | *Klebsiella pneumoniae subspecies pneumoniae* | 24 | S | 22 | S | 15 | R | 8 | R | 42 | S |
|  | 12 | *Salmonella choleraesuis subspecies choleraesuis* | 23 | S | 17 | R | 21 | IR | 21 | S | 29 | S |
|  | 13 | *Yersinia pseudotuberculosis* | 30 | S | 23 | S | 25 | S | 21 | S | 30 | S |
|  | 14 | *Citrobacter freundii* | 50 | S | 50 | S | 50 | S | 50 | S | 50 | S |
|  | 15 | *Klebsiella pneumoniae subspecies ozaenae* | 35 | S | 26 | S | 26 | S | 31 | S | 24 | S |
|  | 16 | *Kluyvera ascorbata* | 25 | S | 14 | R | 23 | S | 19 | IR | 22 | IR |
|  | 17 | *Escherichia coli* | 18 | R | 11 | R | 24 | S | 26 | S | 25 | S |
|  | 18 | *Enterobacter aerogenes* | 42 | S | 43 | S | 30 | S | 40 | S | 39 | S |
|  | 19 | *Salmonella enteritidis* | 39 | S | 31 | S | 25 | S | 32 | S | 20 | R |
|  | 20 | *Klebsiella pneumoniae subspecies pneumoniae* | 20 | IR | 13 | R | 13 | R | 6 | R | 1 | R |
|  | 21 | *Shigella species* | 21 | S | 15 | R | 20 | IR | 18 | IR | 20 | R |
|  | 22 | *Escherichia hermanii* | 30 | S | 28 | S | 25 | S | 21 | S | 24 | S |
|  | 23 | *Kluyvera ascorbata* | 24 | S | 22 | S | 20 | IR | 20 | IR | 21 | R |
|  | 24 | *Klebsiella pneumoniae subspecies pneumoniae* | 50 | S | 50 | S | 50 | S | 50 | S | 50 | S |
|  | 25 | *Shigella flexneri* | 20 | IR | 9 | R | 15 | R | 9 | R | 18 | R |
|  | 26 | *Klebsiella pneumoniae subspecies pneumoniae* | 2 | R | 50 | S | 50 | S | 50 | S | 50 | S |
|  | 27 | *Salmonella enteritidis* | 22 | S | 22 | S | 26 | S | 10 | R | 26 | S |
|  | 28 | *Serratia plymuthica* | 14 | R | 13 | R | 21 | S | 1 | R | 26 | S |
| **YELAHANKA ZONE** | **S. No** | **CLINICALLY RELEVANT** | | | | | | | | | | |
|  | 1 | *Escherichia coli* | 50 | S | 50 | S | 50 | S | 50 | S | 50 | S |
|  | 2 | *Salmonella enteritidis* | 30 | S | 24 | S | 23 | S | 29 | S | 20 | R |
|  | 3 | *Klebsiella pneumoniae subspecies pneumoniae* | 20 | IR | 16 | R | 17 | IR | 20 | IR | 20 | R |
|  | 4 | *Salmonella enteritidis* | 20 | IR | 21 | S | 19 | IR | 15 | R | 16 | R |
|  | 5 | *Enterobacter cloacae* | 23 | S | 15 | R | 21 | S | 14 | R | 12 | R |
|  | 6 | *Salmonella enteritidis* | 20 | IR | 18 | R | 11 | R | 19 | IR | 19 | R |
|  | 7 | *Escherichia coli* | 25 | S | 18 | R | 15 | R | 0 | R | 10 | R |
|  | 8 | *Proteus mirabilis* | 20 | IR | 13 | R | 21 | S | 22 | S | 24 | S |
| **MAHADEVAPURA ZONE** | **S. No** | **CLINICALLY RELEVANT** | | | | | | | | | | |
|  | 1 | *Enterobacter amnigenus* | 20 | IR | 1 | R | 13 | R | 6 | R | 16 | R |
|  | 2 | *Shigella dysenteriae* | 30 | S | 29 | S | 26 | S | 29 | S | 27 | S |
|  | 3 | *Escherichia hermanii* | 31 | S | 27 | IR | 26 | S | 26 | S | 26 | S |
|  | 4 | *Serratia entomophila* | 15 | R | 13 | R | 30 | S | 25 | S | 30 | S |
|  | 5 | *Klebsiella pneumoniae subspecies pneumoniae* | 20 | IR | 13 | R | 22 | S | 15 | R | 17 | R |
|  | 6 | *Proteus mirabilis* | 15 | R | 14 | R | 18 | IR | 29 | S | 16 | R |
|  | 7 | *Yersinia pseudotuberculosis* | 20 | IR | 10 | R | 19 | IR | 28 | S | 19 | R |
|  | 8 | *Pantoea dispersa* | 22 | S | 20 | IR | 20 | IR | 16 | IR | 16 | R |
|  | 9 | *Klebsiella pneumoniae subspecies pneumoniae* | 20 | IR | 22 | S | 18 | IR | 19 | S | 18 | R |
|  | 10 | *Serratia entomophila* | 16 | R | 11 | R | 19 | IR | 12 | R | 18 | R |
|  | 11 | *Klebsiella pneumoniae subspecies ozaenae* | 30 | S | 26 | S | 28 | S | 26 | S | 25 | S |
|  | 12 | *Kluyvera ascorbata* | 30 | S | 23 | S | 25 | S | 15 | R | 35 | S |
|  | 13 | *Klebsiella pneumoniae subspecies pneumoniae* | 18 | R | 18 | R | 17 | IR | 13 | R | 15 | R |
|  | 14 | *Salmonella choleraesuis subspecies choleraesuis* | 18 | R | 8 | R | 22 | S | 18 | IR | 18 | R |
|  | 15 | *Enterobacter amnigenus* | 27 | S | 19 | IR | 30 | S | 23 | S | 24 | IR |
|  | 16 | *Klebsiella pneumoniae subspecies pneumoniae* | 31 | S | 23 | IR | 25 | S | 30 | S | 30 | S |
|  | 17 | *Kluyvera ascorbata* | 25 | S | 19 | IR | 20 | IR | 11 | R | 24 | S |
|  | 18 | *Serratia plymuthica* | 16 | R | 50 | S | 19 | IR | 10 | R | 17 | R |
| **RAJARAJESHWARI NAGAR** | **S. No** | **CLINICALLY RELEVANT** | | | | | | | | | | |
|  | 1 | *Pantoea dispersa* | 21 | S | 18 | R | 7 | R | 25 | S | 19 | R |
|  | 2 | *Proteus mirabilis* | 25 | S | 19 | R | 15 | R | 17 | IR | 1 | R |
|  | 3 | *Klebsiella pneumoniae subspecies ozaenae* | 31 | S | 25 | S | 19 | IR | 20 | S | 30 | IR |
|  | 4 | *Proteus mirabilis* | 25 | S | 2 | R | 26 | S | 9 | R | 11 | R |
|  | 5 | *Pantoea dispersa* | 20 | IR | 15 | R | 17 | R | 14 | R | 15 | R |
|  | 6 | *Salmonella choleraesuis subspecies choleraesuis* | 24 | S | 18 | R | 20 | IR | 18 | IR | 25 | S |
|  | 7 | *Enterobacter amnigenus* | 20 | IR | 15 | R | 21 | S | 1 | R | 17 | R |
|  | 8 | *Salmonella enteritidis* | 15 | R | 8 | R | 20 | IR | 13 | R | 25 | IR |
|  | 9 | *Pantoea dispersa* | 17 | R | 16 | R | 13 | R | 17 | IR | 15 | R |
|  | 10 | *Proteus mirabilis* | 15 | R | 15 | R | 16 | IR | 1 | R | 22 | IR |
| **BOMMANAHALLI** | **S. No** | **CLINICALLY RELEVANT** | | | | | | | | | | |
|  | 1 | *Yersinia pseudotuberculosis* | 25 | S | 33 | S | 26 | S | 33 | S | 30 | S |
|  | 2 | *Citrobacter freundii* | 26 | S | 16 | R | 24 | S | 14 | R | 24 | S |
|  | 3 | *Escherichia hermanii* | 20 | IR | 20 | IR | 15 | R | 25 | S | 16 | R |
|  | 4 | *Klebsiella pneumoniae subspecies pneumoniae* | 23 | S | 17 | R | 25 | S | 26 | S | 20 | R |
|  | 5 | *Citrobacter freundii* | 1 | R | 1 | R | 1 | R | 1 | R | 1 | R |
|  | 6 | *Escherichia coli* | 17 | R | 15 | R | 14 | R | 15 | R | 25 | S |
|  | 7 | *Salmonella enteritidis* | 20 | IR | 15 | R | 20 | IR | 1 | R | 20 | R |
|  | 8 | *Escherichia coli* | 27 | S | 21 | IR | 22 | S | 1 | R | 11 | R |
|  | 9 | *Salmonella choleraesuis subspecies choleraesuis* | 23 | S | 20 | IR | 15 | R | 1 | R | 9 | R |
|  | 10 | *Enterobacter amnigenus* | 30 | S | 39 | S | 45 | S | 39 | S | 40 | S |
|  | 11 | *Shigella species* | 19 | IR | 15 | R | 14 | R | 1 | R | 1 | R |
|  | **S. No** | **ENVIRONMENTALLY RELEVANT** | | | | | | | | | | |
|  | 1 | *Buttiauxella agrestis* | 25 | S | 24 | S | 25 | S | 16 | IR | 26 | S |
| **DASARAHALLI** | **S. No** | **CLINICALLY RELEVANT** | | | | | | | | | | |
|  | 1 | *Escherichia coli* | 20 | IR | 18 | R | 23 | S | 30 | S | 24 | S |
|  | 2 | *Klebsiella pneumoniae subspecies pneumoniae* | 14 | R | 1 | R | 17 | IR | 28 | S | 21 | R |
|  | 3 | *Serratia sp* | 24 | S | 19 | R | 14 | R | 15 | R | 16 | R |
|  | 4 | *Klebsiella pneumoniae subspecies pneumoniae* | 14 | R | 11 | R | 15 | R | 1 | R | 13 | R |
|  | 5 | *Escherichia coli* | 15 | R | 9 | R | 13 | R | 25 | S | 16 | R |
|  | 6 | *Serratia plymuthica* | 25 | S | 19 | IR | 20 | S | 18 | IR | 23 | IR |
