## Supplementary Table 2 for "The First Large-Scale Wastewater Surveillance of AMR to inform Hyperlocal Antibiotic prescribing: A Study from Bengaluru, India"

**Supplementary Table 2:** Non- Enterobacteriaceae isolates classified by Zones of Bengaluru city.

| ZONE | BACTERIA IDENTIFIED | ZONE | BACTERIA IDENTIFIED |
|---|---|---|---|
| SOUTH | CLINICALLY RELEVANT | WEST | CLINICALLY RELEVANT |
| | *Vibrio hollisae* | | *Aeromonas craviae* |
| | ENVIRONMENTALLY RELEVANT | MAHADEVPURA | CLINICALLY RELEVANT |
| | *Pseudomonas fluorescens* | | *Vibrio hollisae* |
| EAST | CLINICALLY RELEVANT | | *Vibrio furnissis* |
| | *Vibrio furnissis* | | *Vibrio hollisae* |
| | *Pseudomonas pseudoalcaligenes* | | *Vibrio furnissis* |
| | *Aeromonas craviae* | | *Vibrio furnissis* |
| | *Vibrio hollisae* | | *Aeromonas craviae* |
| | *Vibrio furnissis* | | ENVIRONMENTALLY RELEVANT |
| | *Vibrio hollisae* | | *Pseudomonas fluorescens* |
| YELAHANKA | CLINICALLY RELEVANT | RAJARAJESWARI | CLINICALLY RELEVANT |
| | *Vibrio furnissis* | | *Vibrio hollisae* |
| | *Pseudomonas areginosa* | | *Vibrio furnissis* |
| | ENVIRONMENTALLY RELEVANT | | *Vibrio furnissis* |
| | *Pseudomonas fluorescens* | | *Vibrio hollisae* |
| BOMMANAHALLI | CLINICALLY RELEVANT | DASARAHALLI | CLINICALLY RELEVANT |
| | *Vibrio fluvialis* | | *Vibrio furnissis* |
| | ENVIRONMENTALLY RELEVANT | | *Vibrio furnissis* |
| | *Pseudomonas fluorescens* | | *Vibrio hollisae* |
