## Supplementary Table 3 for "The First Large-Scale Wastewater Surveillance of AMR to inform Hyperlocal Antibiotic prescribing: A Study from Bengaluru, India"

**Supplementary Table 3:** CLSI guidelines for zone of inhibition for Enterobacteriaceae family

| Zone of inhibition | Resistance (mm) | Intermediate Resistance | Sensitive |
|---|---|---|---|
| Meropenem | <19 | 20-22 | >23 |
| Ertapenem | <18 | 19-21 | >22 |
| Ciprofloxacin | <21 | 22-25 | >26 |
| Cefixime | <15 | 16-18 | >19 |
| Piperacillin/ Tazobactam (TZP) | <17 | 17-19 | >20 |
| CLSI guidelines | | | |
